# Strengthening the health systems at national level for malaria elimination in the Greater Mekong Subregion countries: A qualitative study

**DOI:** 10.1101/2025.09.10.25335540

**Authors:** Win Htike, Win Han Oo, Catherine M. Bennett, Paul A. Agius, Alyssa E. Barry, Freya J. I. Fowkes

## Abstract

Countries in the Greater Mekong Subregion (GMS) have committed to eliminating malaria by 2030. The success of a national malaria programme’s transition from malaria control to elimination is dependent on the readiness of the health system to implement malaria elimination strategies. Understanding the readiness of health systems and what needs to be adapted is key to identifying barriers in achieving malaria elimination goals. To explore the needs for national-level health system readiness, a multi-country qualitative study was conducted in the GMS. Semi-structured interviews were conducted with 39 stakeholders including national malaria policymakers (n=5), basic health staff from Ministries of Health (n=12), managers and field supervisors from malaria implementing partners (n=16) and personnel from technical agencies (n=6). Reflexive thematic analysis of national level health system requirements was carried out aligned with themes adapted from the World Health Organization health system building blocks. Stakeholders discussed barriers in current health system building blocks and areas for improvement needing attention from policymakers and programme implementers. Major barriers identified were the lack of targeted interventions in high-risk groups in national policies, incomplete reporting from private sector, lack of experienced workforce for elimination, administrative constraints in supply chain, declining malaria funding from international donors, and poor compliance to regulations for malaria elimination. To overcome these, national programmes must be equipped with an experienced workforce, sustainable human resource plans, a reliable and smooth procurement and supply chain system, a sustainable funding support, higher-level commitment and a strong technical leadership by the national programme. National programmes must assess national health system readiness for malaria elimination to avoid inefficiencies, financial strain, and unattended gaps, using a systems thinking approach. This study also highlighted the importance of evaluating the national programmes from the perspective of health system needs and readiness for successful transitioning from control to elimination phase.

## Introduction

Countries in the Greater Mekong Subregion (GMS) – Cambodia, Lao People’s Democratic Republic (PDR), Myanmar, Thailand and Vietnam – have committed to eliminating malaria by 2030 [1]. With the exclusion of Myanmar, due to the current political and humanitarian crises, GMS countries have made significant progress in reducing malaria morbidity and mortality [2] and are on track for elimination. To achieve malaria elimination by 2030, each GMS country developed its National Malaria Elimination Plan in the context of its existing health system capacity [1].

National Malaria Elimination Plans of GMS countries [3–7] follow the World Health Organization’s (WHO’s) Global Technical Strategy for Malaria [8], including the importance of early diagnosis and prompt treatment of malaria cases, interruption of transmission by conducting time-bound case and foci investigation and response activities, and strengthening their malaria surveillance systems. Plans highlight universal coverage of malaria prevention and treatment services to high-risk populations, including forest-going mobile and migrant populations, military and construction workers. In addition, they emphasise eliminating vivax malaria, which is complicated by dormant liver stages that can cause relapses. However, radical cure of vivax malaria (elimination of both liver and blood-stages) needs 8-aminoquinolines (primaquine or tafenoquine), which might lead to adverse side effects in patients with glucose-6-phosphate dehydrogenase enzyme (G6PD) deficiency.

Malaria programme reorientation from the control to elimination phase requires adaptation of health system characteristics at national and subnational levels [9, 10]. It is also imperative that National Malaria Elimination Programmes (NMEPs) recognise and formulate malaria elimination strategies comprehensively considering all aspects of the health system [11].

Otherwise, weaknesses in malaria elimination programmes from health system gaps could lead to poor performance and cause delay or failure in achieving malaria elimination goals [12]. This study investigated national-level health system readiness to implement malaria elimination activities across the GMS, providing recommendations for policymakers, higher-level stakeholders, and NMEP managers to evaluate their respective programmes from the health system perspective.

## Materials and methods

### Study setting, design and participants

This study was conducted in four GMS countries (Lao PDR, Myanmar, Thailand and Vietnam) and utilised a descriptive qualitative research approach of key informant interviews and in-depth interviews. The methodological orientation that guided data collection was a phenomenological perspective seeking to understand the participant’s lived experience and knowledge on malaria elimination strategies and activities. Reporting adhered to the consolidated criteria for qualitative research (COREQ) checklist (Supplemental material 1) [13].

A total of 41 malaria stakeholders were approached either in-person or via telephone for qualitative interviews during 31^st^ August 2023 and 11^th^ September 2024; two initially agreed to participate during recruitment but could not be contacted at the time of consent taking.

Study participants were approached purposively based on their role as well as experience and knowledge on malaria elimination programmes and the health system in the GMS, aiming to identify and include potential participants in the study who were information-rich and could provide a full and sophisticated understanding of malaria elimination in the GMS [14]. The sample size and decision to cease recruitment were determined by data saturation.

### Themes

This paper conceptualised the health system from the perspective of WHO health system framework that consists of six core components or building blocks – service delivery; health workforce; health information systems; medical products, vaccines and technologies; healthcare financing; and leadership and governance [15]. Deductive construction of the themes for the qualitative interviews was guided by the conceptual framework on health system readiness dimensions adapted from Colombini *et al* [16] that was conceptualised in the context of malaria elimination in the GMS (Table 1). In order to achieve the goal of malaria elimination, these six inter-connecting building blocks must act harmoniously and in an optimal way, ensuring good access and coverage of malaria services [15].

**Table 1.** Deductive themes to explore needs for health system readiness to implement malaria elimination activities in the Greater Mekong Subregion.

| WHO health system building block | Theme(s) |
| --- | --- |
| Service delivery | • Policy to address universal coverage of malaria services |
| Surveillance | • Responsiveness of surveillance system |
| Elimination workforce | • Long-term human resource plan |
| Products and commodities | • Procurement and supply chain system<br>• Quality control mechanisms |
| Programme financing | • Sustainable plan for malaria elimination |
| Leadership and governance | • Higher-level commitment<br>• Technical guidance from central level |

### Data collection

Interviews were conducted either in-person (n=19) or online (n=19) except one participant who provided their responses in writing due to availability. Interviews were guided by interview topic guides (Supplemental material 2), that were piloted with 5 participants to fine-tune, validate the wording, and estimating the duration of interview. Stakeholders were interviewed by one researcher (WH) in English or Burmese, or in another local language such as Lao or Vietnamese with the assistance of a translator. On average, each interview lasted 60 minutes. No repeat interviews were conducted.

### Data management and analysis

Field notes were taken during interviews by the interviewer. All interviews were audio-recorded using a digital audio recorder, transcribed verbatim, and translated to English, if necessary, except for identifying information of the participants. Transcription and translation were conducted by research assistants and final interview transcripts were independently reviewed and verified for accuracy (WH). Transcripts were not returned to participants for comment.

Reflexive thematic analysis was carried out in six steps – familiarising with the data, generating initial codes, searching for the themes, reviewing the themes, defining and naming themes, and producing the report [17]. Emerging themes during data collection were captured and incorporated into the thematic framework at the data analysis stage. The findings were reported thematically. Key findings were illustrated with direct quotations from the data. Qualitative data analysis was assisted by NVivo software version 15 (licensed to Deakin University).

### Rigour

In this study, rigour was ensured by employing several strategies including purposive sampling, triangulation of qualitative findings with literature during analysis and interpretation of results, ensuring sampling adequacy, and researcher reflexivity from the data collection stage up to the data analysis and reporting stages [18].

To enhance rigour, transcripts of 20% of interviews were randomly reviewed against the original audio recording to ensure accuracy. Two co-investigators (WH and WHO) analysed the transcripts independently and the team compared and discussed findings until consensus on themes was achieved. Lastly, an audit trail was maintained during the coding process to ensure all analyses could be traced back to the original data.

### Positionality statement

Although the researchers have extensive experience working in malaria elimination programmes and health system strengthening in GMS, none has experience in government service in the GMS and the understanding is external for government programmes.

### Ethical considerations

The study protocol was approved by the Alfred Ethics Committee (199/23), Deakin University Human Research Ethics Committee (2023-233), National Ethics Committee for Health Research of Lao PDR (19/NECHR), and Institutional Review Board of Vietnam (25-2023/HDDD). Written informed consent was obtained from all study participants (Supplemental material 3). Strict measures were taken to ensure privacy, respect, confidentiality and dignity of all participants. Data collection was conducted in private locations where both the visual and audial privacies were ensured.

### Inclusivity in global research

Additional information regarding the ethical, cultural, and scientific considerations specific to inclusivity in global research is included in the Supporting Information (Supplemental material 4).

## Results

In this analysis, 39 stakeholders were interviewed including national level malaria policymakers and programme managers (n=5), field level supervisors and basic health staff from Ministries of Health and NMEPs (n=12), managers and field supervisors from malaria implementing partners (n=16) and personnel from technical agencies and research organisations (n=6). Mean age of stakeholders was 45 years (standard deviation, SD: 10 years) with an average experience of 13 years in the malaria field (SD: 10 years), and mostly responsible for field implementation and management (62%, 24/39) (Supplemental material 5, Additional table 1). Stakeholders discussed the health system requirements at the national level for malaria elimination, which are presented as per the themes adapted from the WHO health system building blocks.

### Service delivery

Stakeholders emphasised that universal access to malaria prevention, diagnosis and treatment must be ensured when entering the elimination phase. Malaria prevention and treatment services should not only target the general population but also focus on high-risk populations and high-transmission areas to ensure that residual transmission is effectively interrupted. Guided interviews with stakeholders revealed a gap at the national level regarding service delivery policies among high-risk migrants in the GMS. Stakeholders believed that current mainstay vector control measures in the GMS (long lasting insecticidal nets (LLINs) and targeted indoor residual spraying) are not sufficient to protect the residual transmission among these high-risk populations. They mentioned that personal protective measures are yet to be included in the national policies and used in combination with existing interventions to effectively reduce residual transmission among migrants. To explain this, stakeholders described an ideal scenario where mobile and migrant people could sleep under the LLIN while they were at home and use repellent and insecticide-treated clothing when they went into the forest.

> “*In the situations where LLINs and indoor residual spraying are used, they are effective, and they are applied indoors… in Asia Pacific, we have a big problem in that many of our primary vectors are outdoor biting… if they bite outdoors, they are not exposed to our primary tools used globally, which are LLIN and indoor residual spraying.*” (A regional level key informant and vector control specialist)

Moreover, stakeholders from different levels believed that G6PD testing could improve treatment adherence and contribute significantly to radical cure of vivax malaria cases. Currently, all the countries in the GMS except Myanmar recommend mandatory G6PD testing before prescribing primaquine or tafenoquine. However, stakeholders advised that the adherence to this recommendation was questionable in some areas due to unavailability of test kits, and willingness and skill of health staff to perform G6PD testing due to the complex nature of the testing.

> *“… they (health staff) don’t care about the G6PD testing because they had not experienced any side effects of primaquine… but we need to strengthen G6PD testing to improve treatment adherence (for vivax cases).”* (A field supervisor)

### Surveillance

In elimination settings, the national malaria surveillance system must capture all cases detected at different settings such as health facilities, general hospitals, community provider (e.g. village health volunteers), private clinics and hospitals, and other institutions (e.g. military, forestry, labour). However, stakeholders discussed that it was challenging to collect data from the private sector and military in some GMS countries. Stakeholders believed that such challenge was partly due to lack of proactive dialogue at national level with authorities from those sectors by NMEP. At the national level, NMEP should lead the dialogue between different actors to advocate and coordinate to include them in the malaria surveillance system.

> *“Even though there is a law to notify all malaria cases, we don’t receive malaria reports from military. I think the Ministry of Health needs to improve coordination with them (military).”* (A field supervisor)

In addition to the case-based epidemiological surveillance system, the NMEPs also need to establish and strengthen other surveillance systems in the malaria elimination programme such as drug resistance marker monitoring and therapeutic efficacy studies, entomological and insecticide resistance marker surveillance and susceptibility assays. Even though countries in the GMS have been implementing these surveillance measures, stakeholders expressed their concern that the technical capacity of health staff needs to be strengthened to get accurate and reliable information.

> *“For entomology, the local Centre of Disease Control team needs to do different kinds of surveys. They could do vector species identification, but not insecticide resistance and susceptibility tests.”* (A field supervisor)

### Elimination workforce

NMEP programme managers and leaders need to have strong technical knowledge and experience on malaria control and elimination in addition to efficient programme management skills. During interviews, stakeholders reported that malaria elimination was a technically challenging process, and that NMEPs needed a more experienced workforce especially at the national level to navigate discussions with other departments within and beyond the Ministry of Health, to orientate the programme in the right direction, and to lead the programme with a systems thinking approach.

> “*When it comes to this last mile (of malaria elimination), you need the most experienced hands…*” (A regional level key informant)

However, in the GMS, stakeholders reported that most of the senior staff with experience and knowledge of malaria control and elimination, and technical capabilities such as surveillance or resistance monitoring were either retired or reassigned to other disease programmes before achieving elimination. To retain an experienced workforce and equip the programme for the successful implementation of malaria elimination activities, stakeholders recommended that the NMEPs advocate the Ministry of Health to develop a long-term human resource plan.

> *“Until it (malaria) is eradicated, you will need to keep that competence about malaria in your workforce… They (Ministries of Health) need to ask (themselves) how to maintain that tropical medicine capacity within their workforce, even if it is a rare disease.”* (A regional level key informant)

### Products and commodities

During interviews, stakeholders shared that the forecasting and quantification must be realistic as well as responsive. For example, one of the GMS countries had forecasted its malaria cases in a downward trend until 2030 and used those figures to quantify the commodity requirement. However, the number of malaria cases had increased recently in that country and the NMEP was not willing to revise or amend the previous forecast that no longer reflected the needs of the programme. The key informant made an educated guess that the country would be unavoidably facing the issue of stock-out in a near future.

Moreover, stakeholders suggested that procurement of malaria commodities should be smooth and swift. Stakeholders discussed that when malaria burden is declining, countries usually need fewer commodities such as antimalarial drugs. In the GMS, it was challenging for each NMEP to undertake the procurement by themselves because of the small quantity of products needed in each country. To resolve the situation, the principal recipient of the donor (the main organisation responsible for implementing and managing a malaria grant) organised the procurement process on behalf of the NMEPs, and it was noted that the process was faster and easier than when the NMEPs did it by themselves.

To elaborate on the importance of a smooth supply chain system, stakeholders discussed an example from one country which had experienced changes in the supply chain. In the past, different implementing partners needed to submit their commodity requirement to the principal recipient of the donor, from which they received their quota and distributed to the respective field operation sites. However, the logistic supply chain system in that country is now centralised and controlled by the NMEP. Implementing partners could not issue their required commodities from the principal recipient anymore. Instead, they need to submit commodity requests to the NMEP and sub-national health authorities, which is challenging due to bureaucratic and complex administrative red tape. As a result, most of the implementing partners faced the unavoidable issue of stock-outs at service delivery points. Stakeholders made their point that the supply chain system in an elimination setting should be swift and smooth regardless of being centralised or not.

Furthermore, proper quality assurance and control mechanisms should be in place at the national level to ensure that the malaria commodities meet their expected quality and standards. Currently, countries in the GMS are procuring WHO pre-qualified products, which also undergo routine quality assurance and control measures before distributing to the service delivery points. Post-marketing quality surveillance is also done for malaria RDTs, medicines and LLINs. Stakeholders suggested that these regular quality assurance and control mechanisms should be sustained over time and strengthened if required.

> “*Regular quality assurance and control are not only for rapid diagnostic tests (RDTs) and antimalarials, but also for LLINs, insecticides, and other laboratory equipment and reagents.”* (A national level key informant)

Some stakeholders expressed their opinion that antimalarials should be widely available in different sectors. In the GMS region, malaria testing and treatment services are provided free of charge at public health facilities. However, facilities are not accessible to every patient. Some might choose to attend private clinics and then need to purchase their medicines from pharmacies. Stakeholders mentioned that a significant number of pharmacies no longer stock antimalarials due to a decline in demand from the general public. Since the national malaria supply chain systems in the GMS does not cover the private sector, some stakeholders suggested that quality antimalarials should also be available in the private sector. However, some stakeholders also expressed their concerns regarding weak regulation against fake and substandard drugs and artemisinin monotherapies in the market even though they are officially banned in GMS countries.

### Programme financing

Stakeholders unanimously agreed that intensive implementation of malaria elimination programmes costs more than that of a control programme. What is more, international funding for malaria programme has been redirected to high transmission areas in Africa as the malaria burden decreased in the GMS. With the declining malaria burden in the GMS, malaria was not a priority for higher-level policymakers and the government reduced the budget for malaria programmes. This was particularly alarming given malaria elimination needs sustainable and adequate funding.

> *“I don’t know if you’ve seen that… Where almost (malaria) elimination had occurred, people (policymakers) took their foot off the accelerator (giving example of driving a car), and malaria resurged.”* (A regional level key informant)

It was clear from the interviews that all GMS countries relied heavily on international donors for malaria elimination given domestic funding is minimal. As per the discussion of one regional level key informant, Sri Lanka was close to malaria elimination in late 1990s but experienced a massive resurgence of malaria cases due to government funding cuts for malaria elimination programmes. Stakeholders suggested that NMEPs should think about sustainable solutions to reduce the operational costs and formulate strategies to cope with the decline in international and domestic funding for malaria elimination.

As a short-to medium-term solution, stakeholders suggested a close collaboration between NMEP and civil society organisations to ensure effective utilisation of available resources from international donors. A suggested long-term solution for sustainable malaria elimination was to integrate malaria services into primary healthcare services so that all the disease elimination programmes could share the existing domestic resources and infrastructure. In addition, stakeholders also suggested other innovative solutions such as introducing malaria into health insurance schemes and collecting taxation from commercial companies and earmarking it for malaria elimination.

> “*And it’s especially in those stages, elimination and post-elimination, where countries need to absolutely prioritise and make very sure that they have adequate resources for domestic funding of these programmes … domestic funding…., internal resources are critically important for countries to maintain and wean themselves off international donors as far as possible.*” (A regional level key informant)

### Leadership and governance

Stakeholders unanimously agreed that higher-level political commitment and engagement are extremely important for achieving malaria elimination. Many stakeholders believed that malaria elimination could not be achieved with standalone efforts from the NMEPs and needed coordination and cooperation with other departments and ministries, otherwise there could be bottlenecks or delays to field activities. Leaders of GMS countries committed to eliminating malaria by 2030 and stakeholders suggested that NMEPs should continue advocating their respective governments for a sustained higher-level commitment and support regarding malaria elimination.

At the national level, the NMEPs must ensure that regulations and standing orders related to malaria elimination were strictly followed by all concerned parties at different levels. Stakeholders also mentioned that the NMEPs needed to issue standard operating procedures so that there was no discrepancy in bringing these regulations and standing orders into action. Each GMS country has issued various regulations relating to malaria elimination (e.g. mandatory case notification, banning of artemisinin monotherapies), however, stakeholders from some countries expressed that there were challenges in following them.

> “*Following the regulation for mandatory malaria case notification is not smooth and easy in practice since there is no standard operating procedure or specific direction from the national programme. As a result, malaria cases from private sector are neither reported nor notified to the public health authorities till now.*” (A field supervisor)

Stakeholders also expressed the important role of research in malaria elimination. They mentioned that research was needed to find new and innovative methods and approaches, and to evaluate existing strategies, interventions and tools, while transitioning into an elimination phase. Many stakeholders agreed that research projects should be in line with the needs of the NMEPs. Stakeholders also suggested that research uptake by policymakers and higher-level stakeholders should be improved. As suggested by the stakeholders, NMEPs needed to organise multidisciplinary workshops involving both programme and academic stakeholders to develop a research agenda, share the research findings and explore ways to gain policy impact from the research findings.

> “*In the short term, research findings often face challenges in being integrated into control and elimination programmes… But that research should and must continue. Otherwise, we’re going to sit with challenges that all tools become ineffective, and we don’t have new tools to replace them with.*” (A regional level key informant and researcher)

Even though GMS countries developed standard operating procedures (SOPs) and guidelines on malaria elimination, stakeholders believed that field implementation at the lower levels would not be effective and efficient without strong technical leadership and guidance from NMEPs. They mentioned that NMEPs should keep (malaria elimination) procedures clear, concise and easy to understand (for the implementers), otherwise there could be discrepancies between different organisations in implementing malaria elimination strategies at field levels. Furthermore, stakeholders suggested developing simple, clear and user-friendly standardised forms and SOPs for data quality assurance and good documentation practices according to the national level strategic needs for transforming malaria surveillance into a core intervention.

> “*The forms for foci investigation and responses are too complex and lengthy. They are technically sound, but not practical for us to use. Only the VBDC team leader could fill it out and not easy (to complete) for other staff, so most of the forms come back (from the township levels to the regional level) empty.*” (A field supervisor)

## Discussion

Malaria elimination is a resource-intensive time-bound intervention that could only be built upon an infrastructure of a strong national health system. This study highlighted that malaria elimination needs inputs from all six WHO health system building blocks at the national level. Stakeholders identified certain national-level gaps in the existing health system building blocks such as insufficient targeted tools and approaches for migrants, difficulty in collecting malaria data from private sector and military, lack of experienced staff to lead the elimination team, complicated and lengthy supply chain, declining funding landscape and poor compliance to regulations that needed attention from NMEP managers and national level policymakers. Some of these gaps are becoming more challenging as case numbers decline. If malaria elimination in the GMS is to prevail, the national-level health system needs to be strengthened across all six WHO health system building blocks as summarised in Table 2.

**Table 2.**
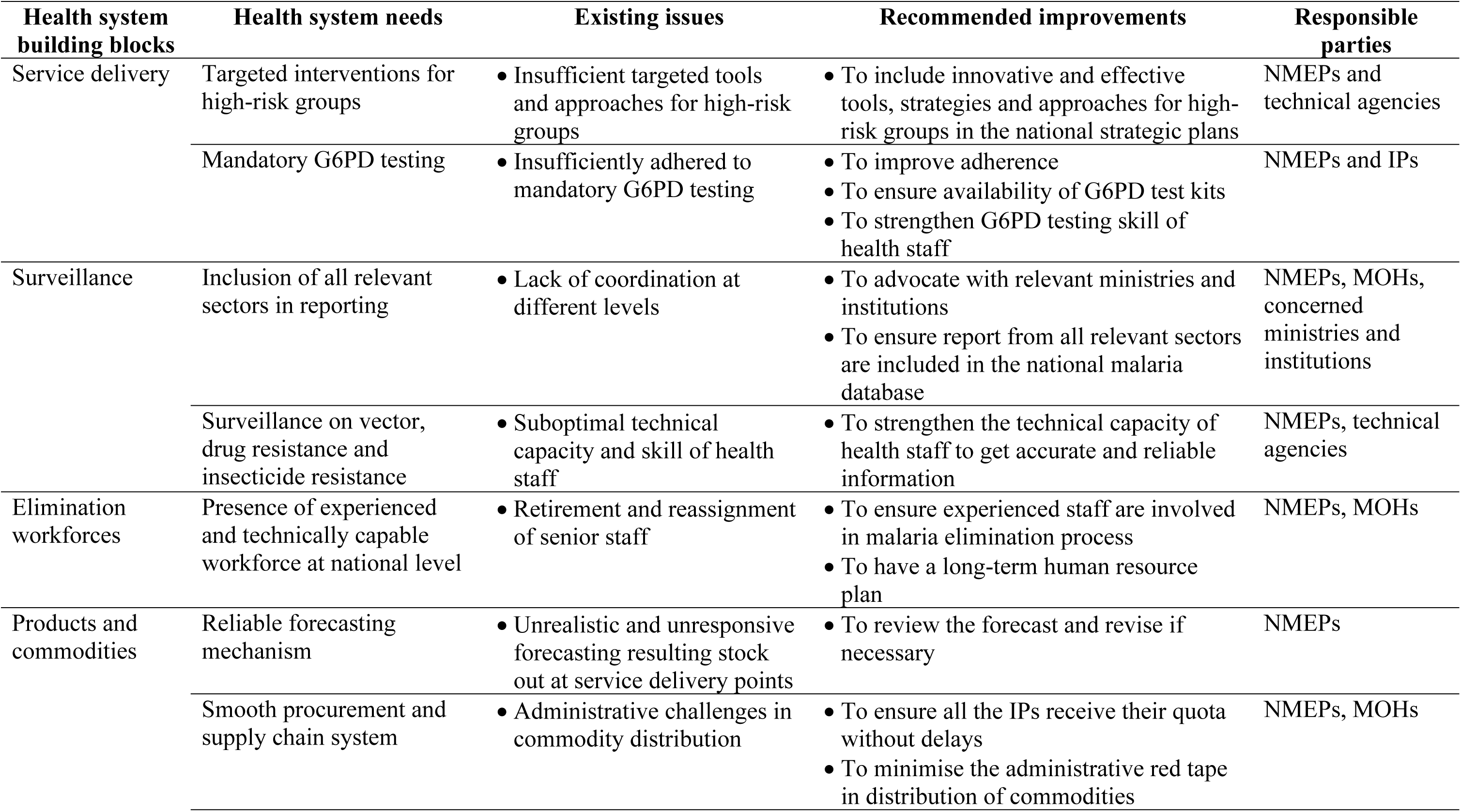
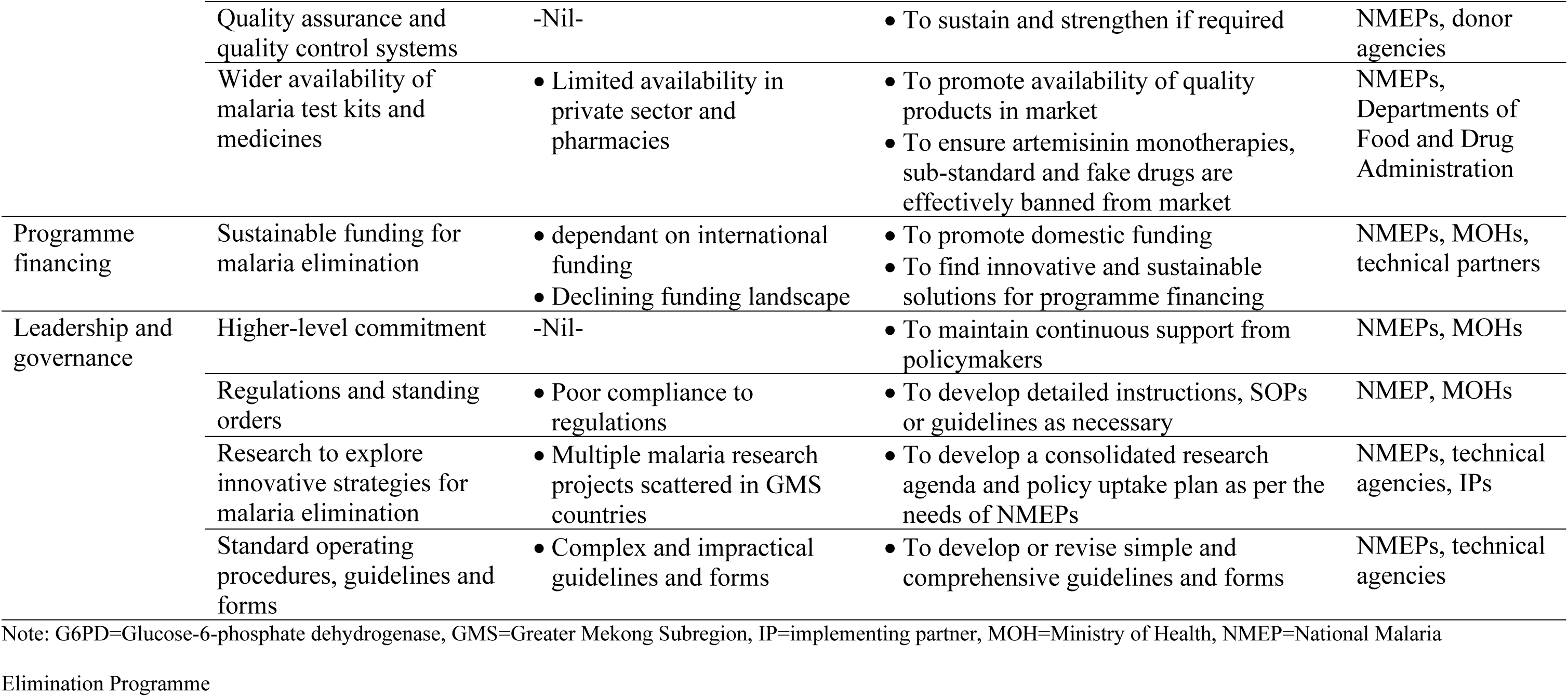
National level health system needs and strengthening for malaria elimination in Greater Mekong Subregion.

| **Health system building blocks** | **Health system needs** | **Existing issues** | **Recommended improvements** | **Responsible parties** |
| --- | --- | --- | --- | --- |
| Service delivery | Targeted interventions for high-risk groups | • Insufficient targeted tools and approaches for high-risk groups | • To include innovative and effective tools, strategies and approaches for high-risk groups in the national strategic plans | NMEPs and technical agencies |
|  | Mandatory G6PD testing | • Insufficiently adhered to mandatory G6PD testing | • To improve adherence<br>• To ensure availability of G6PD test kits<br>• To strengthen G6PD testing skill of health staff | NMEPs and IPs |
| Surveillance | Inclusion of all relevant sectors in reporting | • Lack of coordination at different levels | • To advocate with relevant ministries and institutions<br>• To ensure report from all relevant sectors are included in the national malaria database | NMEPs, MOHs, concerned ministries and institutions |
|  | Surveillance on vector, drug resistance and insecticide resistance | • Suboptimal technical capacity and skill of health staff | • To strengthen the technical capacity of health staff to get accurate and reliable information | NMEPs, technical agencies |
| Elimination workforces | Presence of experienced and technically capable workforce at national level | • Retirement and reassignment of senior staff | • To ensure experienced staff are involved in malaria elimination process<br>• To have a long-term human resource plan | NMEPs, MOHs |
| Products and commodities | Reliable forecasting mechanism | • Unrealistic and unresponsive forecasting resulting stock out at service delivery points | • To review the forecast and revise if necessary | NMEPs |
|  | Smooth procurement and supply chain system | • Administrative challenges in commodity distribution | • To ensure all the IPs receive their quota without delays<br>• To minimise the administrative red tape in distribution of commodities | NMEPs, MOHs |
|  | Quality assurance and quality control systems | -Nil- | • To sustain and strengthen if required | NMEPs, donor agencies |
|  | Wider availability of malaria test kits and medicines | • Limited availability in private sector and pharmacies | • To promote availability of quality products in market<br>• To ensure artemisinin monotherapies, sub-standard and fake drugs are effectively banned from market | NMEPs, Departments of Food and Drug Administration |
| Programme financing | Sustainable funding for malaria elimination | • dependant on international funding<br>• Declining funding landscape | • To promote domestic funding<br>• To find innovative and sustainable solutions for programme financing | NMEPs, MOHs, technical partners |
| Leadership and governance | Higher-level commitment | -Nil- | • To maintain continuous support from policymakers | NMEPs, MOHs |
|  | Regulations and standing orders | • Poor compliance to regulations | • To develop detailed instructions, SOPs or guidelines as necessary | NMEP, MOHs |
|  | Research to explore innovative strategies for malaria elimination | • Multiple malaria research projects scattered in GMS countries | • To develop a consolidated research agenda and policy uptake plan as per the needs of NMEPs | NMEPs, technical agencies, IPs |
|  | Standard operating procedures, guidelines and forms | • Complex and impractical guidelines and forms | • To develop or revise simple and comprehensive guidelines and forms | NMEPs, technical agencies |

Note: G6PD=Glucose-6-phosphate dehydrogenase, GMS=Greater Mekong Subregion, IP=implementing partner, MOH=Ministry of Health, NMEP=National Malaria
Elimination Programme

Health system building blocks are interconnected and bottlenecks in a building block could have negative impacts on others. For instance, NMEPs need strong commitment from the federal government for long-term funding, and other support to progress towards malaria elimination [19, 20]. Lack of higher-level commitment and support could result in decreased support for malaria elimination activities in terms of budget, human resources, and materials and commodities, leading to disruptions in delivering malaria services and delays in undertaking reactive surveillance and response activities [21, 22]. Needless to say, onward malaria transmission might not be interrupted effectively in such situations and the country’s goal of malaria elimination could be jeopardised.

Moreover, GMS countries need changes in national level policy and guidelines to bring about effective implementation at the service delivery level. As indicated by the WHO framework for malaria elimination, universal access to malaria prevention, diagnosis and treatment needs to be ensured when entering the elimination phase. Malaria prevention and treatment services should not merely target the general population, but also the high-risk populations and high-transmission areas to make sure residual transmission has been cut off [11]. GMS countries have taken certain measures to interrupt the onward transmission of malaria among high-risk groups, but the mainstay interventions for malaria prevention and vector control did not prevent the majority of infective bites by dominant outdoor vectors [23]. Previous studies have proven the effectiveness of topical repellent in reducing malaria infections either as a standalone intervention or as part of a personal protection package [24–26], however, NMEPs still need to find a way to effectively incorporate such strategic and essential vector control interventions to address residual transmission among high-risk populations.

National authorities in all GMS countries have committed to eliminating malaria by 2030 [27]. The NMEPs have formulated and implemented national strategies and plans to achieve this ambitious goal of malaria elimination [4–7, 28]. However, the health system must have the ability to rapidly and sustainably adapt these policies, processes and infrastructure to support integration of malaria elimination activities on top of existing control interventions [16, 29, 30].

When transitioning from control to elimination, the NMEP must ensure all the health systems needs are in place to host the elimination activities. Otherwise, the lack of preparedness and readiness can result in inefficient use of resources, causing financial strain and potentially undermining public trust in health systems [31]. Furthermore, without proper evaluation on health system readiness, critical gaps in the health system may go unnoticed, hindering the overall success of elimination efforts [32]. The NMEP managers need to practice a systems thinking approach and it is important to assess the readiness of health system at the national level for malaria elimination to identify and address issues and bottlenecks to optimise the country’s elimination efforts.

### Strengths and limitations of the study

This study provided a higher-level view on health system needs and strategies to strengthen national health systems for malaria elimination in the GMS since it included a range of malaria stakeholders with different expertise and backgrounds. Reflexive strategies were applied throughout the study to improve the rigour and trustworthiness of the research so that the research findings are credible and accurately reflect the stakeholders’ experiences and perspectives.

However, due to administrative constraints, the study did not include some participant groups in GMS such as stakeholders from Cambodia, government staff from Thailand and representatives from military. Furthermore, the study did not include grassroots level staff such as field workers and community volunteers although their voice could provide community perspectives of health system strengthening at a national level. It would be beneficial to include a complete set of study population groups to ensure study findings are applicable for the entire GMS region.

## Conclusions

As recommended by the WHO, the choice of malaria elimination strategies and interventions should not only be based on transmission intensity but also on operational capacity and health system readiness. Programme managers and relevant stakeholders should evaluate the National Malaria Programmes from the perspective of health system needs and readiness for successful transitioning from control to elimination phase. Otherwise, it could result in untimely transitioning to elimination phase, unrealistic milestone and target settings, and sub-optimal and inefficient resource utilisation. Moreover, the gaps in the health system should be identified and addressed in due course to avoid delays in achieving malaria elimination by 2030.

## List of abbreviations

G6PD: glucose-6-phosphate dehydrogenase enzyme
GMS: Greater Mekong Subregion
Lao: PDR Lao People’s Democratic Republic
LLIN: long-lasting insecticidal net
NMEP: National Malaria Elimination Programme
RDT: rapid diagnostic test
WHO: World Health Organization

## Contributors

WH conceptualised and designed the study under the guidance of FJIF, AB, CB, PAA and WHO. WH conducted the interviews and performed the qualitative data analysis under supervision of WHO, who also reviewed the themes identified during the analysis process. WH prepared the first draft of the manuscript; and all authors reviewed and contributed to the final manuscript. All authors approved and read the final manuscript.

## Acknowledgements

We would like to thank the National Malaria Elimination Programmes of Lao PDR, Myanmar, Thailand, and Vietnam as well as local health authorities for administrative support. We would also like to extend our thanks to Burnet Institute staff Chad Hughes, Julie Tartaggia and Phone Myint Win, and Health Poverty Action staff Wang Bangyuan, Thet Lynn, Than Vu and Ngo Thi Van Anh for coordination and management support.

## Data availability

Data used and/or analysed during the current study are available from the corresponding author on reasonable request.

## Supporting information

Supplemental material 1: Completed consolidated criteria for reporting qualitative research (COREQ) checklist

Supplemental material 2: Interview topic guides

Supplemental material 3: Sample of participant information and consent form

Supplemental material 4: Inclusivity in global research checklist

Supplemental material 5: Additional table

